# Survival Analysis on Papillary Thyroid Cancer Through Cox Proportional Hazards Model

**DOI:** 10.64898/2025.12.23.25342949

**Authors:** Dilmi Abeywardana, Malinda Iluppangama, Chris P. Tsokos

## Abstract

Cancer accounts for one in six deaths globally, making patient survivorship critical for medical professionals. Thyroid cancer incidence is rising in the United States, with Papillary Thyroid Cancer (PTC) representing approximately 80% of cases. This study develops a semi-parametric Cox Proportional Hazards model to investigate associations between PTC patient survival time and five risk factors: age, sex, malignant tumor size, race, and cancer stage. Our final Cox-PH model, which satisfies all necessary assumptions, identified three statistically significant individual risk factors and three two-way interaction terms that influence mortality hazard rates. We ranked all factors and their interactions by hazard ratio to determine the strongest contributors to patient mortality. These findings provide insights into how demographic and clinical characteristics affect PTC prognosis, potentially informing risk stratification and treatment planning for this increasingly prevalent cancer type.

## 1 Introduction

One of the leading causes of death in today’s world is cancer, which accounts for one out of six deaths. In 2022, the number of new cancer cases was about 20 million, and 9.7 million deaths from cancer globally. Moreover, new cancer cases are predicted to be 35 million by 2050, which is a 77% increase compared to 2022. The most challenging issue with cancer is that approximately 400,000 children are diagnosed with it yearly [28] [21]. This increasing trend of cancer patients is an important factor, and many research studies have been done based on identifying risk factors and their contribution to the survival of cancer patients.

Thyroid cancer is the 12th leading cancer in the world and the seventh most common malignancy in women. Recently, the new rate of thyroid cancer has been growing rapidly compared to other cancers in the United States. In 2025, the most recent estimates of the American Cancer Society for Thyroid cancer are about 44,020 new cases of Thyroid cancer, of which 12,670 are men and 31,350 are women, and about 2,290 deaths due to Thyroid cancer, of which 1,090 are men and 1,200 are women, in the United States. In addition, Thyroid cancer is one of the cancers that is diagnosed at a younger age, with an average age of 51 years [24] [14].

There are four main Thyroid cancer types: Papillary, Follicular, Medullary, and Anaplastic. Papillary Thyroid Cancer (PTC), also known as Papillary Carcinoma, is the most common type of Thyroid cancer, accounting for 8 out of 10 cases of Thyroid cancer. PTC grows slowly and develops in only one lobe of the thyroid gland; however, sometimes it spreads to lymph nodes in the neck. It can be successfully treated with a high survival rate. Follicular Thyroid cancer is the second most common type, accounting for 1 in 10 Thyroid cancer cases. However, Medullary Thyroid cancer accounts for less than 5% of Thyroid cancers, while Anaplastic Thyroid cancer is rare, with 2% of Thyroid cancers. [27] [20]. Thus, this study focuses on the most common type of Thyroid cancer, that is, patients with **PTC**. In our previous study, we developed a real data-driven analytical model to predict the malignant tumor size of a PTC patient and identified the optimal values of risk factors that minimize the malignant tumor size of a PTC patient. This work will help develop new treatment strategies to improve the clinical decision-making process and patient’s survival outcomes [22] [2].

Thyroid cancer starts when a cell begins to lose control in the thyroid gland, producing hormones that help regulate metabolism, heart rate, blood pressure, and body temperature. In front of the neck, the thyroid cartilage, called Adam’s apple, contains a thyroid gland that cannot be seen or felt by most people. The thyroid gland has a butterfly shape with two lobes, called left and right, which are joined by a narrow piece of tissue called an isthmus. The size of a healthy thyroid can be a little larger than a quarter [27] [15].

A lump (nodule), voice change(including hoarseness), fatigue, cough without cold, difficulty swallowing, swollen lymph nodes in the neck, and pain in the neck and throat are the most common symptoms of Thyroid cancer. This type of cancer may be caused by exposure to radiation as a child or at a young age due to medical treatments, radiation fallout from nuclear weapons or power plant accidents, the amount of iodine in the diet or the family history of thyroid cancer in a parent or sibling [20] [3] [18] [16] [12] [4]. According to the literature, Thyroid cancer can develop at any age but is more common in people at age 30s to 60s. The diagnosis of Thyroid cancer peaks earlier for women, often in their 40s and 50s, than for men, usually in their 60s and 70s. In addition, Thyroid cancer occurs about three times more often in women than in men [25] [6] [10]. The authors discussed the relationship of the size of the malignant tumor with the other risk factors of PTC patients in their previous research papers [7]. The medical professional performs additional tests to identify the growth of cancer and establishes the stage of the patient’s cancer, which is an indication of the severity of the cancer by representing how far the cancer has spread outside of the thyroid [26].

When it comes to cancer, the most worrying factor is the survivability of the patient. The most commonly applied technique for survival analysis is the non-parametric Kaplan-Meier (KM) estimation, which has been applied to analyze the survival of patients in our previous study [8]. KM estimation is a univariate analysis that describes survival under one factor (survival time), ignoring the effect of other risk factors related to cancer disease. Thus, the Semi-parametric Cox Proportional Hazards (Cox-PH) model is an alternative to the KM method by simultaneously addressing the effect of several risk factors on the survival time.

The purpose of our study is to identify how the survival time of patients with PTC is affected based on several covariates, including the patient’s Age, Malignant tumor size, Sex, Stage of cancer, and Race through the Cox Proportional Hazards (Cox-PH) model. The Cox-PH model is widely used in medical research to address the hazard ratio of a patient with respect to his or her selected risk factors. Provides the association between the survival time of a patient and risk factors to identify whether a particular risk factor affects the patient’s survival positively or negatively, and what are the most influential risk factors for survivorship. Our final Cox-PH model includes five individual covariates and three two-way interactions, but only three individual covariates (Sex, Malignant tumor size, and Race) are statistically significant, and the other two covariates (Age and Stage) are associated with statistically significant interactions. This proposed Cox-PH model has satisfied all key assumptions by validating the accuracy and high quality of the model. In our previous work, we have applied the Cox-PH approach to analyze the survival of patients who underwent the treatment, deep brain stimulation for Parkinson’s Disease and help to identify the factors that will significantly affect to the survival of patients with Parkinson’s disease and help to develop strategic personalized treatments to imporve survival outcomes of the patients [13]. In this chapter, Section 2 includes details of the data used to develop the Cox-PH model.

Section 3 discusses the methodology for the Cox-PH model and its diagnosis. In Section 4, we discuss the development process of the Cox-PH model, including our final proposed model. The validation of the high-quality model by satisfying all necessary assumptions is described in Section 5. Finally, Section 6 summarizes the final contribution of our study.

## 2 The Real Data

In this study, we have utilized the data obtained from the National Cancer Institute’s (NCI’s) Surveillance, Epidemiology, and End Results (SEER) database [1]. The original data consists of a large number of patients, and it has been cleaned by selecting patients whose cause of death is Thyroid cancer and by removing missing observations. Thus, our final sample consists of 315 PTC patients. The most important variable for survival analysis is the survival times of the patients, which is the response variable in months for our study. There are five risk factors: Age, Malignant tumor size, Sex, Stage of cancer, and Race that have been used to identify the relationship to the survival rate of PTC patients through the Cox-PH model. The data description and data network of the covariates are given below, in Figure 1.

1. Age (*X*_1_): Age of the patient when he/she was diagnosed with PTC.

- If age is greater than or equal to 20 and less than or equal to 70 = 0
- If age is greater than or equal to 71 and less than or equal to 96 = 1
2. Tumor Size (*X*_2_): Malignant tumor size of the patient when he/she was diagnosed with PTC, measured in millimeters (mm).
3. Sex (*X*_3_): Gender of the patient.

- Male = 0
- Female = 1
4. Stage (*X*_4_): SEER database has three main stages based on how far the Thyroid cancer has spread.

- Localized = 0: The cancer has not spread outside of the thyroid.
- Regional = 1: The cancer has spread outside of the thyroid to nearby structures.
- Distant = 2: The cancer has spread to distant parts of the body, such as bones.
5. Race (*X*_5_): PTC patient is either a white person or other.

- White = 0
- Other = 1

**Figure 1:**
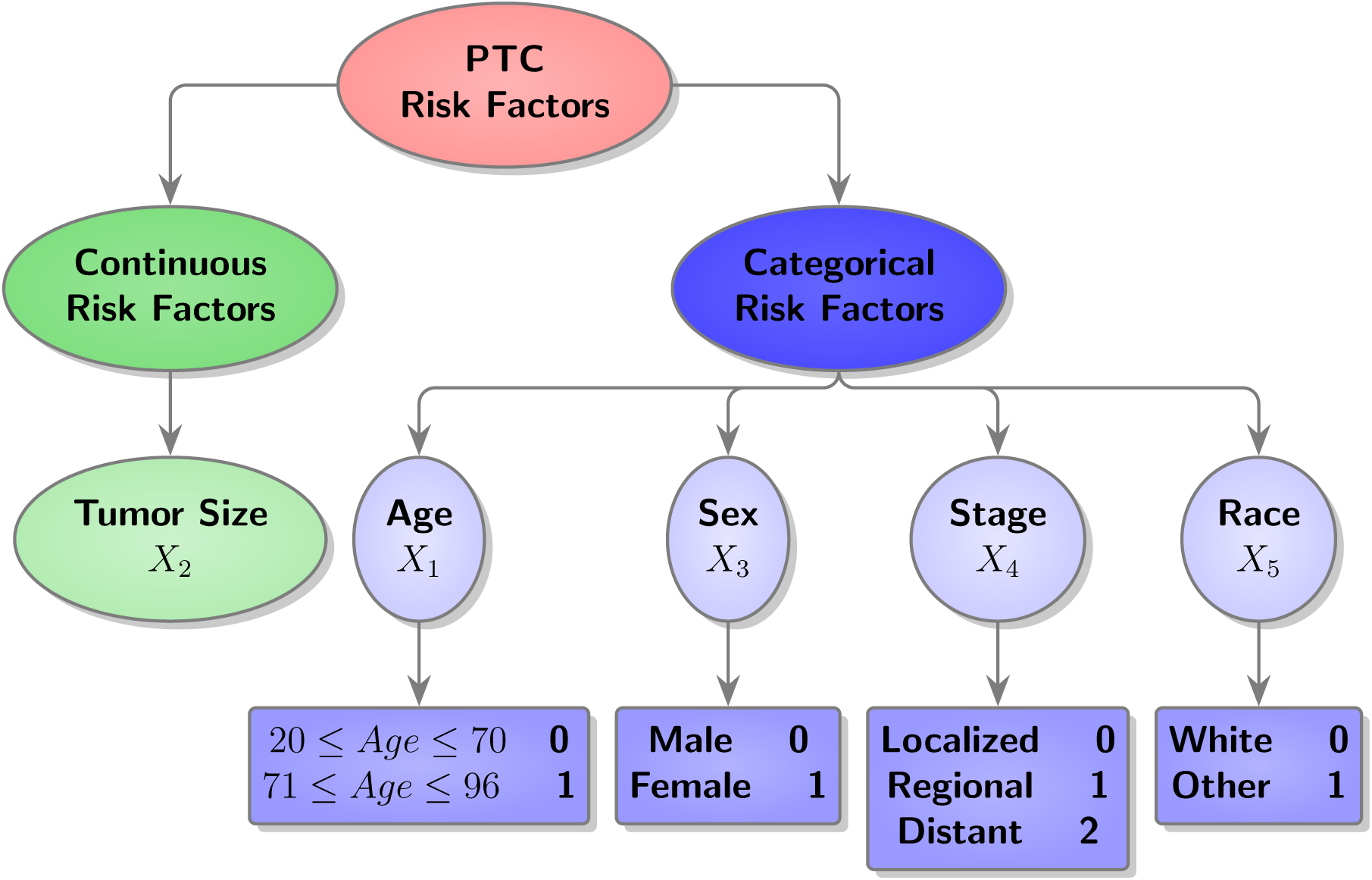
Data Network of PTC Risk Factors

## 3 Methodology

### 3.1 Cox Proportional Hazards Model

Cox proportional hazards, Cox-PH model, is the most commonly used semi-parametric regression method in survival analysis that was introduced by D. R. Cox in 1972 [9]. The Cox-PH model is a semi-parametric method because it combines the parametric and non-parametric parts and doesn’t require any assumption about the probability distribution of the event (death) times. The non-parametric part is used to model the hazard function, and the parametric part is used to model the relationship between the hazard function and covariates through regression tools for the Cox-PH model or accelerated failure time models. The Cox-PH model is commonly used in medical research to identify the association between the time to event (death) and the covariates (risk factors).

Let *T* be a random event time, then the probability that an event occurs prior or at time *t* (probability of failure by time *t* ) is the cumulative distribution function (CDF), given by,

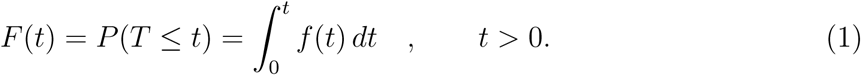

Then the probability that an individual has survived after time *t* is the survival function, given by,

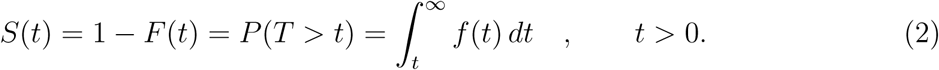

The hazard function is the instantaneous risk of an event at time *t*. That is, given that an individual has not experienced the event prior to time *t*, the probability of the event occurring at the next subsequent time starting at time *t* per unit time is the hazard function, which is denoted by,

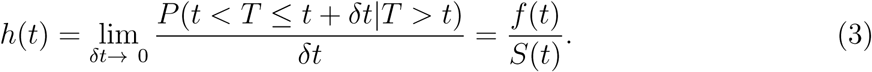

Then the cumulative hazard function is derived by,

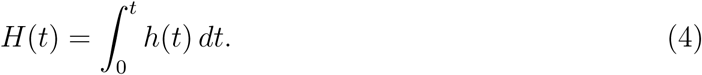

Let *h_i_*(*t*) denote the hazard function at time *t* for an individual. Then the Cox-PH model can be written as,

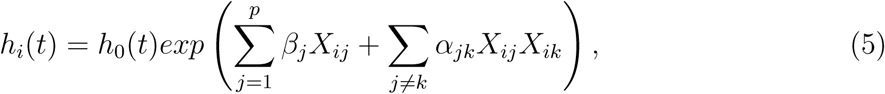

where *t* is the survival time, *β_j_*is the coefficient parameter for *j^th^*covariate and *α_jk_*is the coefficient parameter for the interaction between *j^th^* and *k^th^* covariates. The *h*_0_(*t*) is the arbitrary baseline hazard function, and it equals the hazard function when all the covariates are equal to zero. Thus, the baseline hazard function doesn’t depend on time *t*. Moreover, there is no intercept term for the final Cox-PH model.

We use partial likelihood estimation to estimate the parameters of the Cox-PH model given in equation 5 because it is a combination of parametric and non-parametric parts.

Then the linear component of the Cox-PH model in equation 5, can be expressed as follows,

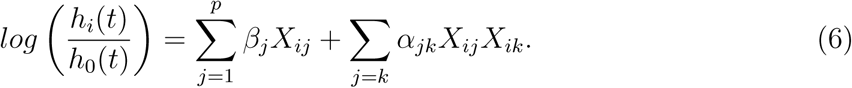

Thus, in the Cox-PH model, the hazard ratio, HR (exp(coef)), compares the hazard of the event (in our case, death) at time *t* between two individuals with *X* = *x* vs. *X* = *x* + 1 if the covariate is continuous or between two individuals with specific levels if the covariate is categorical. That is, for a single continuous covariate, *X* = *Tumorsize* (measured in mm), HR gives the relative risk of the event for an individual who has a tumor size 1 mm greater than another individual. Thus, the hazard ratio for any two individuals z and *z*^∗^ is independent of time *t* and can be expressed by,

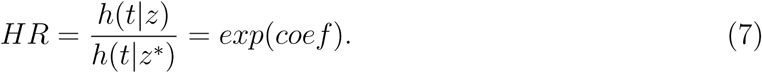

Thus, HR measures the relative risk while *β* gives the log of that relative risk. Three important facts can be derived from the HR. They are,

- If *HR >* 1(*β >* 0), then the covariate is positively associated with event probability. (Negatively associated with survival), i.e., an indication of a bad prognostic factor.
- If *HR <* 1(*β <* 0), then the covariate is negatively associated with event probability. (Positively associated with survival), i.e., an indication of a good prognostic factor.
- If HR *≈* 1, then there is no effect from the covariate. i.e., an indication of a neutralized prognostic factor.

Further readings on the subject area can be found in the literature [9] [19] [30] [5] [23].

### 3.2 Diagnosis for Cox-PH Model

The final Cox-PH model should satisfy the following three key assumptions given below to be a robust model.

1. Should follow the proportional hazard assumption.
2. Should be free from outliers or influential observations.
3. Should have linear relationships between the log hazard and continuous covariates.

There are a few commonly used residual methods to validate the above assumptions for the Cox-PH model. The Schoenfeld residuals can be used to validate the proportional hazard assumption. We can visualize the Martingale residuals to identify the nonlinearity among the log hazard and the continuous covariates. The Deviance residuals illustrate the influential observations or outliers in the final Cox-PH model. We discuss each residual method in more detail below.

1. **Proportional hazards assumption** The scaled Schoenfeld residuals can be used to check one of the main assumptions on the Cox-PH model, the proportional hazards assumption. The proportional hazards assumption has a non-significant relationship between the residuals and the time. This can be tested through a statistical test, using the R package cox.zph(). The test shouldn’t be statistically significant for all covariates of the Cox-PH model, and it has an additional test, called global, which considers the model as a whole and cannot be statistically significant to validate the proportional hazards assumption. Also, we can approach the graphical visualization of scaled Schoenfeld residuals, and the random pattern against time (independent of time) is an indication of the proportional hazards assumption.
2. **Linearity of continuous covariates** We need to satisfy the assumption that the continuous covariates address the linearity, and T. Therneau, P. Grambsch, and T. Fleming [29], have introduced a graphical visualization of the Martingale residuals against the continuous covariates with the fitted lowess smoothing (locally weighted scatter-plot smoothing) to assess the linear form of them. A representation of a pattern is an indication of a violation of the linearity assumption. This linearity assumption should be satisfied by all continuous covariates; however, the categorical variables don’t have any effect on linearity.
3. **Deviance residuals / Dfbeta values** The deviance residuals can be obtained through a normalized transformation of martingale residuals, and the residuals should be symmetric around zero with a standard deviation of one. A very small or large value on the deviance residuals plot is an indication of an influential observation (or an outlier). Additionally, we can plot dfbeta values, which provide the estimated changes in the regression coefficients by removing each observation in turn to identify influential points (or outliers). A high value of dfbeta might be an influential observation, and we should investigate such observations more deeply.

Further readings of this subject matter can be found in the literature [11] [29] [17].

In the next section, we proceed to develop a Cox-PH model for the survival times of PTC patients having five individual risk factors and their two-way interactions, and confirm the adequacy and the quality of the resulting model by validating all assumptions.

## 4 Cox-PH Model Developing Process

We develop a Cox-PH model that takes into consideration the relationship between covariates (risk factors) and the hazard function of the PTC patients. While developing the Cox-PH model, it is important to satisfy the proportional hazards assumption, which assumes that there is a proportional relationship between the hazard function and different levels of the covariates. Thus, as an initial graphical approach, we can plot *log*(*−log*(*S*^^^(*t*))) vs. time, which is the log of the transformed KM estimate of the survival function vs. time for each significant categorical variable, to illustrate the parallelism within the levels. Moreover, after the model has been developed, we can apply the test discussed in Section 3.2 along with the graphical approach.

In our case, initially, the covariate, Age, is continuous, but after the model was built, it didn’t satisfy the proportional hazards assumption. Thus, we applied the remedy, called stratification, on the variable Age and divided it into two groups as described in Section 2, and this method may lead to more valid estimates for the covariates. Figure 2, given below, illustrates the parallelism of the survival curves of the age groups.

**Figure 2:**
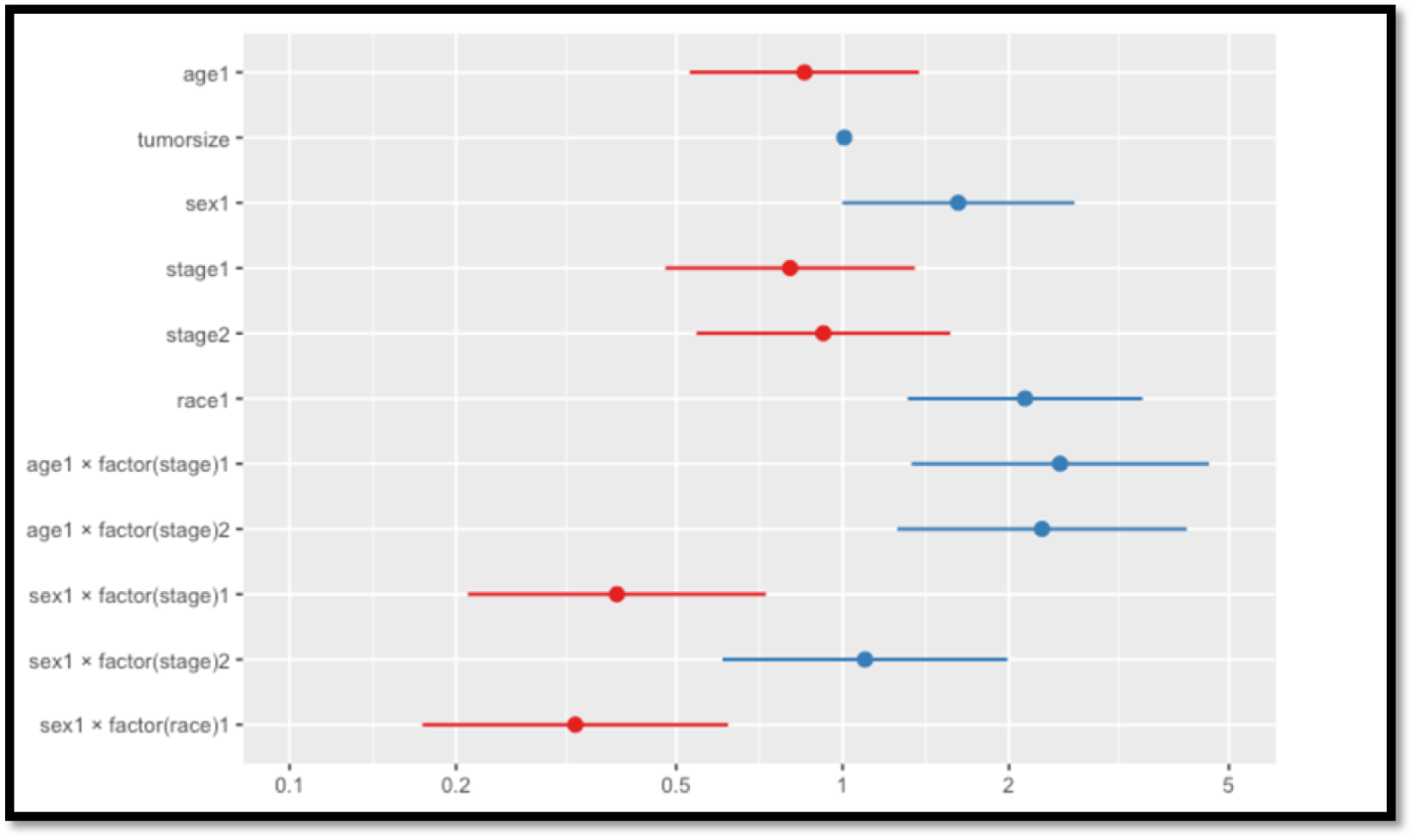
Survival Curves for the Two Age Groups

First, we utilized the stepwise model building procedure with Akaike Information Criterion, AIC (*AIC* = *−*2*logL*+2*p* where *LogL* is the log-likelihood function and p is the number of parameters) to build our initial Cox-PH model, including five individual risk factors together with their two-way interactions (5C2 = 10). The stepwise variable selection method is based on both forward selection and backward elimination steps. Then, the backward elimination method was applied on the remaining terms to remove insignificant terms (p-value *>* 0.05) from the model one by one and reached the final Cox-PH model with only statistically significant individual terms and their two-way interactions at a 5% level of significance. Thus, the proposed Cox-PH model for the survival times of PTC patients is statistically significant with five individual risk factors (only three covariates are individually statistically significant, and two covariates are associated with significant interactions) and three two-way interactions, given by,

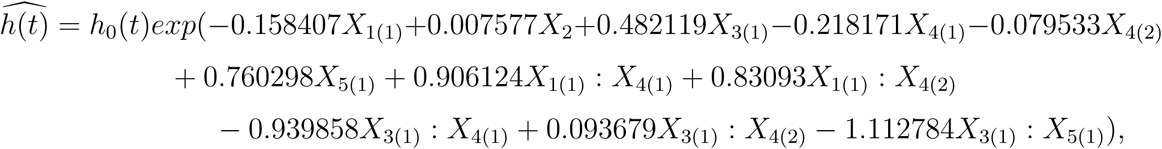

where *h(t)* is the hazard function at time *t*, *h*_0_(*t*) is the arbitrary baseline hazard function, *X_i_*_(*l*)_ is the *i^th^* covariate with level *l* (for a categorical risk factor) as described in Section 2.

Then the linear relationship between the survival of the PTC patients and the covariates (risk factors) can be rearranged as follows,

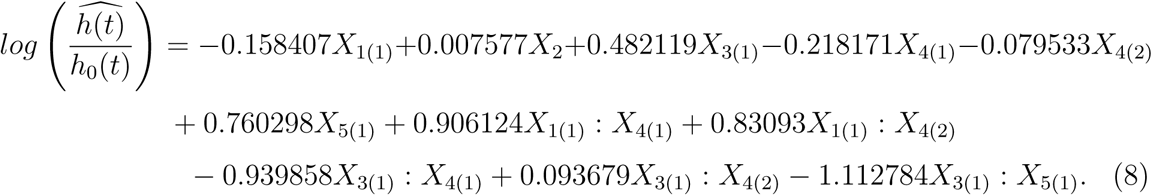

Table 1, below, contains the information about our proposed Cox-PH model with the estimates of model coefficients (parameters), hazard ratios (exp(coef)), standard errors of coefficients, and 95% lower and upper confidence intervals for the hazard ratios. Figure 3 visualizes the 95% confidence intervals of the hazard ratios, corresponding to the terms that are included in our final Cox-PH model. Moreover, Table 2, below, provides the global statistical significance values for the overall significance of the proposed Cox-PH model by using three hypothesis tests: Likelihood ratio test, Wald test, and Score test, and all p-values (*<* 0.05) confirm that the proposed Cox-PH model is statistically significant at the 5% level of significance. These three tests are asymptotically equivalent, but, for small samples, the likelihood ratio test slightly outperforms the Wald test, and both outperform the Score test. In our case, the sample size is large enough to have equivalent results for three tests as given in Table 2.

**Figure 3:**
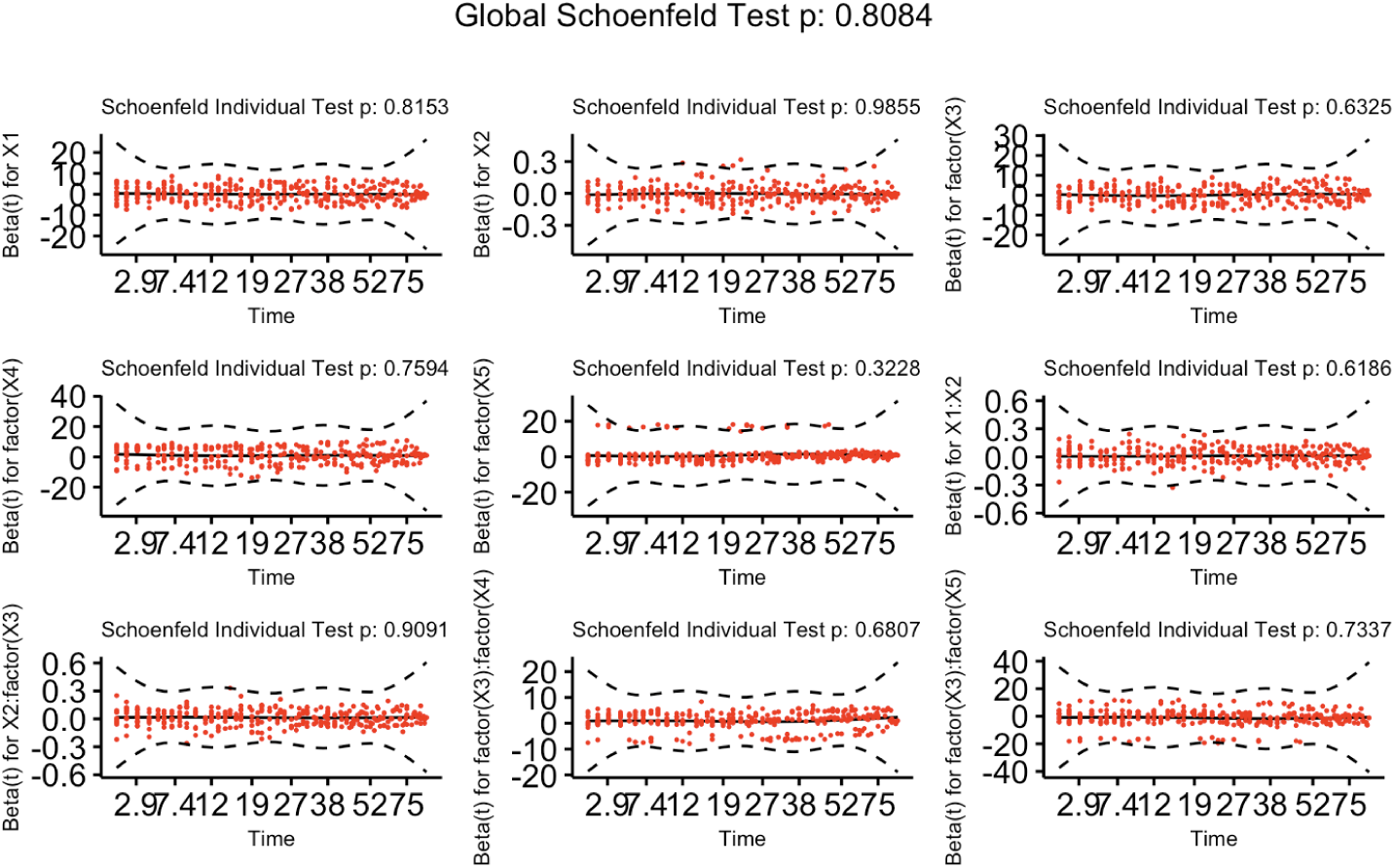
95% Confidence Intervals for the Terms in the Proposed Cox-PH Model

**Table 1:** Coefficients and Hazard Ratios of the Proposed Cox-PH Model.

| Covariate | Coefficient | Hazard Ratio ($\exp(\text{coef})$) | SE(coef) | 95% Lower | 95% Upper |
| --- | --- | --- | --- | --- | --- |
| $X_{1(1)}$ | -0.158407 | 0.853503 | 0.243495 | 0.5296 | 1.3755 |
| $X_2$ | 0.007577 | 1.007606 | 0.002574 | 1.0025 | 1.0127 |
| $X_{3(1)}$ | 0.482119 | 1.619503 | 0.246937 | 0.9981 | 2.6277 |
| $X_{4(1)}$ | -0.218171 | 0.803988 | 0.265130 | 0.4782 | 1.3518 |
| $X_{4(2)}$ | -0.079533 | 0.923548 | 0.269788 | 0.5443 | 1.5671 |
| $X_{5(1)}$ | 0.760298 | 2.138914 | 0.249634 | 1.3113 | 3.4888 |
| $X_{1(1)} : X_{4(1)}$ | 0.906124 | 2.474711 | 0.316173 | 1.3317 | 4.5989 |
| $X_{1(1)} : X_{4(2)}$ | 0.830930 | 2.295453 | 0.307767 | 1.2557 | 4.1960 |
| $X_{3(1)} : X_{4(1)}$ | -0.939858 | 0.390683 | 0.316553 | 0.2101 | 0.7266 |
| $X_{3(1)} : X_{4(2)}$ | 0.093679 | 1.098207 | 0.302765 | 0.6067 | 1.9879 |
| $X_{3(1)} : X_{5(1)}$ | -1.112784 | 0.328643 | 0.324658 | 0.1739 | 0.6210 |

**Table 2:** Global Statistical Significance of the Proposed Cox-PH Model.

| Hypothesis Test | P-value |
| --- | --- |
| Likelihood ratio test | 9e-13 |
| Wald test | 2e-12 |
| Score test | 2e-13 |

As we discussed in Section 3.1, the hazard ratio *(exp(coef))* measures the relative risk. If the coefficient is positive (*β*^^^ *>* 0), i.e., hazard ratio, *(exp(coef)) >* 1, then the event of occurrence is high with a bad prognostic factor. However, if the coefficient is negative (*β*^^^ *<* 0), i.e., hazard ratio, *(exp(coef)) <* 1, then the event of occurrence is low with a good prognostic factor. In our case, the coefficient of Sex (*β*^^^_3_ = 0.482119 *>* 0) is positive, and it means a female PTC patient has a higher risk for death with a bad prognostic while a male PTC patient has a lower risk of death at any time. Moreover, the covariate, Race, has a positive coefficient indicating that a patient from another race has a higher risk of death compared to a white patient. However, the interactions between Age and Stages have positive coefficients, indicating a higher risk for the PTC patients at an age between 70 - 96 in regional and distant stages compared to the patients at an age between 20 - 70 in the localized stage. Similarly, all significant terms in the proposed Cox-PH model can be interpreted.

In addition, we proceed to rank the individual covariates and their two-way interactions of our proposed statistically significant Cox-PH model based on the prognostic effect on the survival times of PTC patients by using the hazard ratios *(exp(coef))*, as given in Table 3, below.

**Table 3:** Ranking of the Contributing Covariates and Interactions Using Hazard Ratios.

| Covariate | Hazard Ratio $(exp(coef))$ | Rank |
| --- | --- | --- |
| $X_{1(1)} : X_{4(1)}$ | 2.474711 | 1 |
| $X_{1(1)} : X_{4(2)}$ | 2.295453 | 2 |
| $X_{5(1)}$ | 2.138914 | 3 |
| $X_{3(1)}$ | 1.619503 | 4 |
| $X_{3(1)} : X_{4(2)}$ | 1.098207 | 5 |
| $X_2$ | 1.007606 | 6 |
| $X_{4(2)}$ | 0.923548 | 7 |
| $X_{1(1)}$ | 0.853503 | 8 |
| $X_{4(1)}$ | 0.803988 | 9 |
| $X_{3(1)} : X_{4(1)}$ | 0.390683 | 10 |
| $X_{3(1)} : X_{5(1)}$ | 0.328643 | 11 |

According to the ranking of the significant contributing covariates and their interactions based on hazard ratios, the first two most contributing factors for the survival of PTC patients are the interactions between the Age and Stages; however, they are not individually significant. Thus, they are ranked at seven, eight, and nine, respectively. Race is the individual significant covariate with the highest contributing risk factor for the survival of PTC patients, following Sex and Malignant tumor size.

## 5 Validation of the Cox-PH Model

Three key assumptions should be satisfied to have a robust Cox-PH model as outlined in Section 3.2. In this section, we proceed to verify those three assumptions.

### 5.1 Test the proportional hazards assumption

We can apply the following formal statistical test to justify whether the proposed Cox-PH model satisfies the proportional hazards assumption. In Table 4, all p-values are greater than 0.05. Thus, the test is not statistically significant for each of the covariates, and the global test is also not statistically significant, confirming that our proposed Cox-PH model satisfies the proportional hazards assumption.

**Table 4:** P-values for Testing the Proportional Hazards Assumption.

| Covariate | P-value |
| --- | --- |
| $X_1$ | 0.72 |
| $X_2$ | 0.60 |
| $X_3$ | 0.92 |
| $X_4$ | 0.72 |
| $X_5$ | 0.34 |
| $X_1 : X_4$ | 0.75 |
| $X_3 : X_4$ | 0.83 |
| $X_3 : X_5$ | 0.82 |
| Global | 0.81 |

Additionally, the graphical representation of scaled Schoenfeld residuals against time for all individual covariates (risk factors) and two-way interactions in our proposed Cox-PH model can be used to test the proportional hazards assumption, and scaled Schoenfeld residuals should be independent of time, and a plot with a non-zero slope is an indication of the violation of the assumption. The plots of scaled Schoenfeld residuals for our fitted Cox-PH model are given in Figure 4, below.

**Figure 4:**
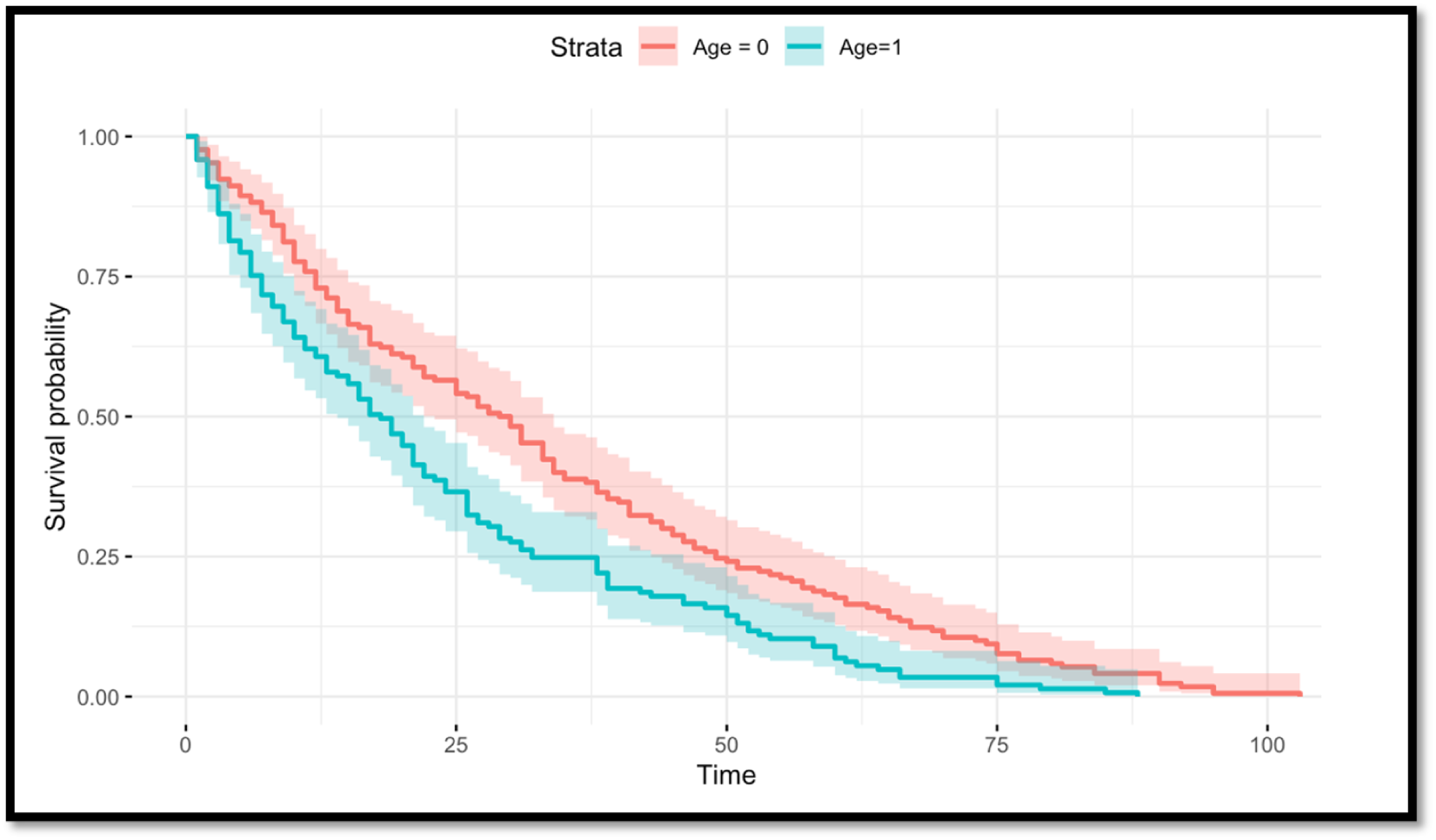
Plots of Scaled Schoenfeld Residuals for Each Term of the Proposed Cox-PH Model

In Figure 4, all the plots of Schoenfeld residuals follow a zero slope. Thus, there is no violation of the proportional hazards assumption, and scaled Schoenfeld residuals are independent of time.

### 5.2 Check for the influential (or outlier) observations

The deviance residuals can be used to identify the influential/ outlier observations. The deviance residual is a normalized transform of the martingale residual and is roughly symmetrically distributed about zero with a standard deviation of 1. In our case, there are no major influential/ outlier points in the deviance residuals plot of the final Cox-PH model, as shown by Figure 5, below.

**Figure 5:**
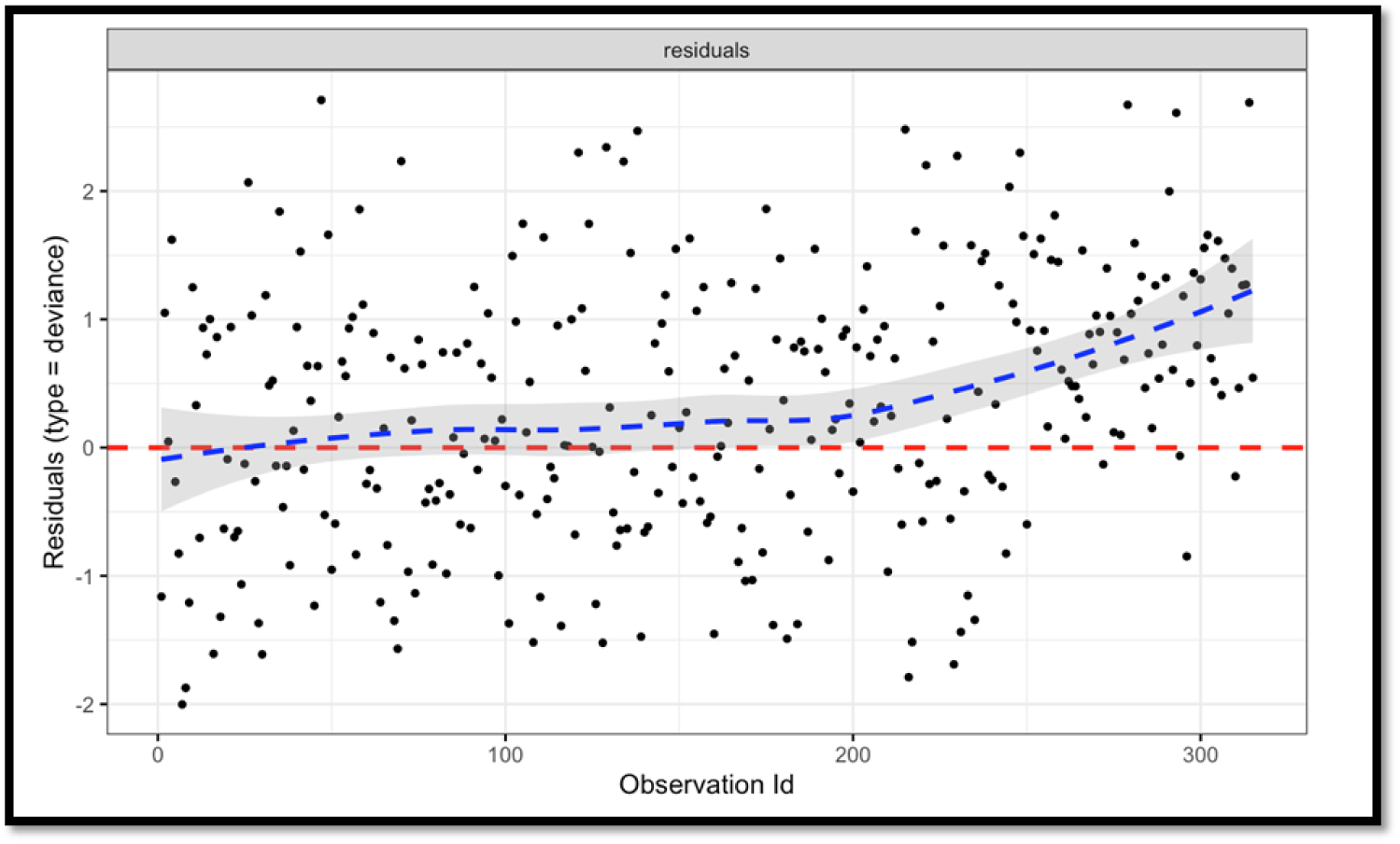
Deviance Residuals Plot for the Proposed Cox-PH Model Additionally, we have graphically visualized the dfbeta values whose estimates influence the observations on a Cox-PH model. Figure 6, below, shows that there are no high values for dfbeta. Thus, it confirms no influential/ outlier observations in our proposed Cox-PH model.

### 5.3 Testing linear functional form of Continuous variables

Continuous covariates are expected to follow a linear functional form. Martingale residuals vs. continuous covariates can be used to detect non-linearity. In our case, we have only one continuous covariate, the Malignant tumor size, and Figure 7, represents the linear form of that covariate using Martingale residuals, and satisfies the assumption of linearity.

**Figure 6:**
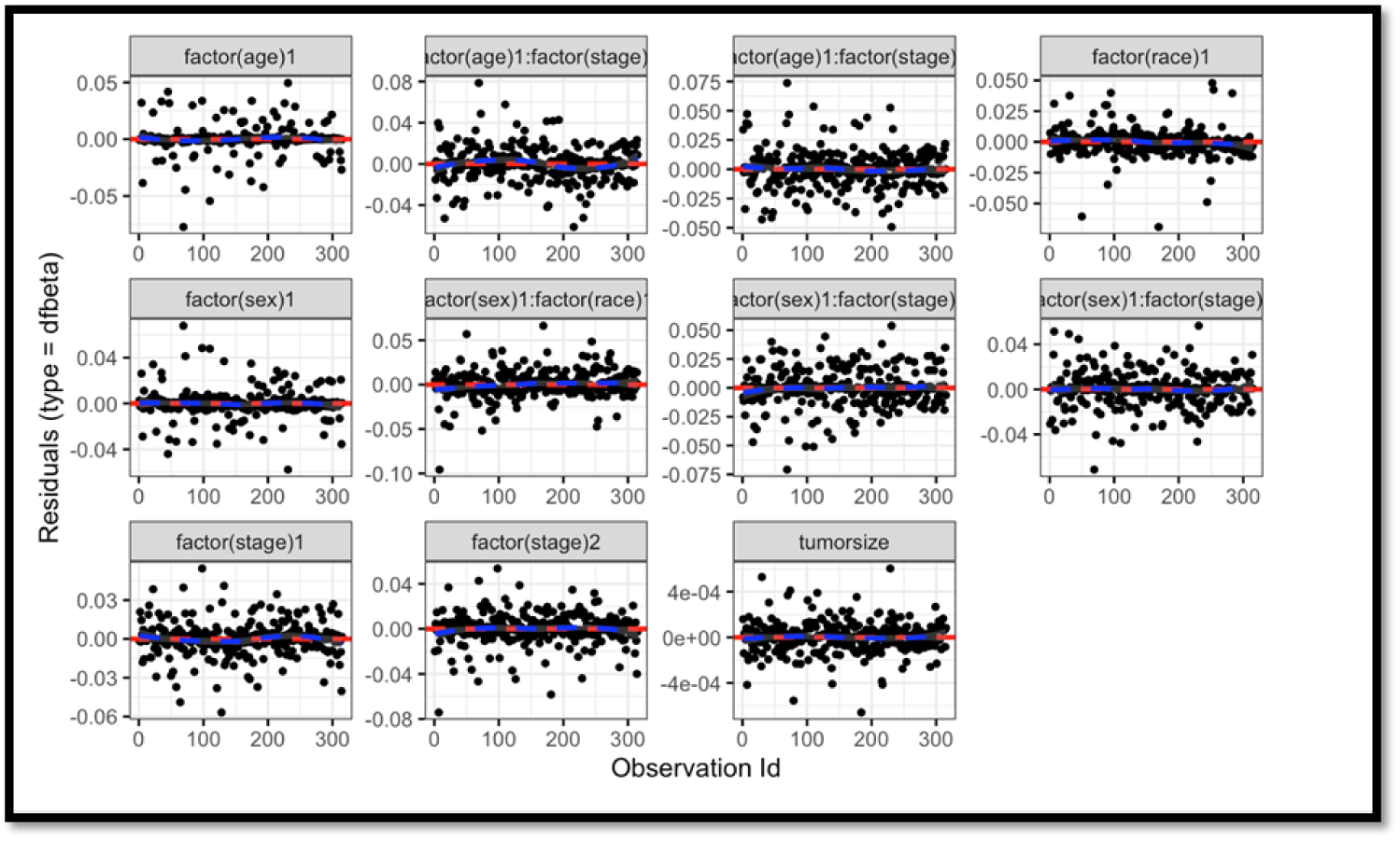
Dfbeta Values for the Proposed Cox-PH Model

**Figure 7:**
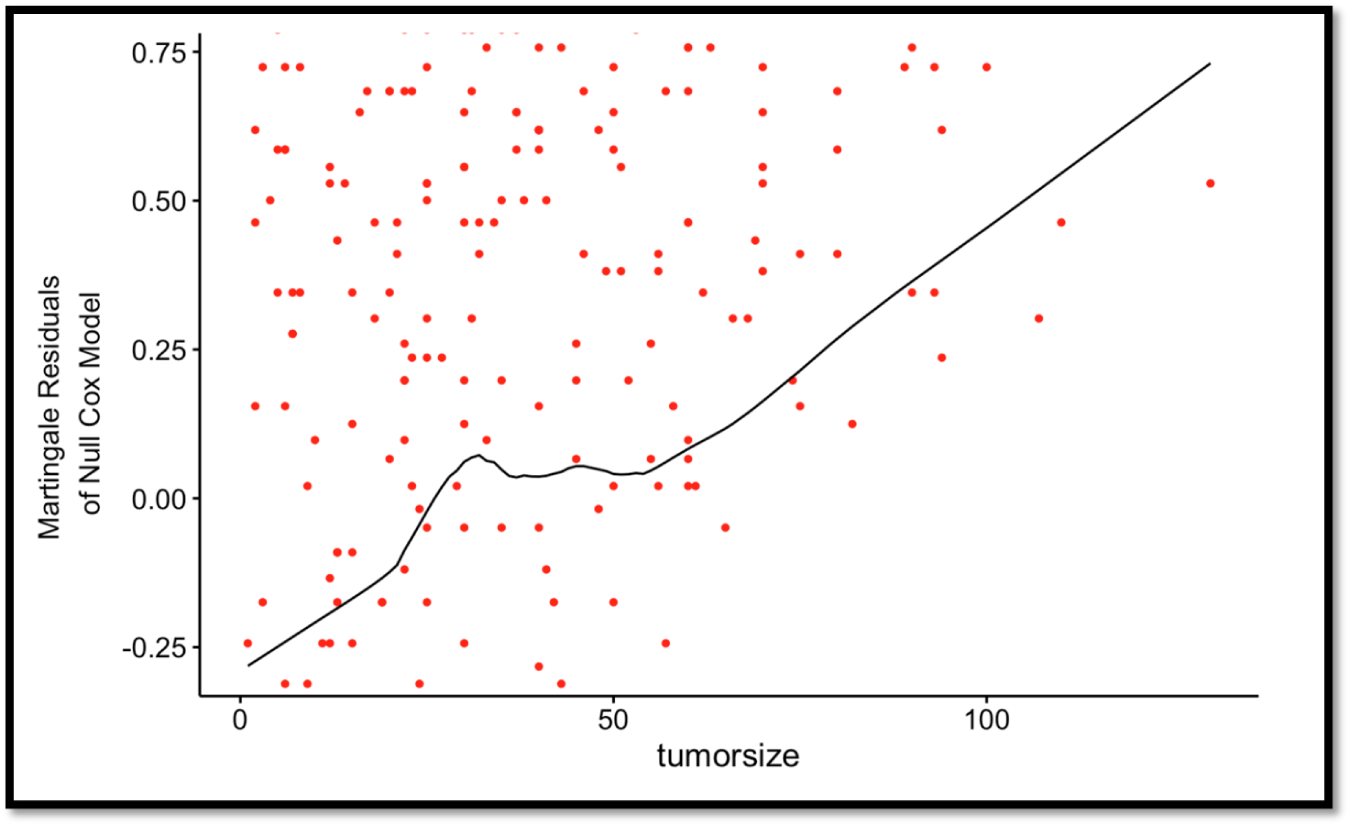
Martingale Residuals Plot for the Continuous Variable

Since the three key assumptions are satisfied by our proposed Cox-PH model, it is robust and accurately characterizes the association between the survival of the PTC patients and the individual covariates and their two-way interactions.

We proceed to visualize the Cox-Snell residuals to examine the overall fit of our proposed Cox-PH model for PTC patients.

### 5.4 Cox-Snell residuals

The Cox-Snell residuals taken from a complete data set should follow a unit exponential distribution, and it works more robustly with larger samples of observations. This can be used to examine the overall fit of the Cox-PH model. If the Cox-Snell residuals follow a straight line with a zero intercept and unit slope, then the fitted Cox-PH model confirms that the subject model is a good fit. For our final Cox-PH model, the Cox-Snell residuals result in a straight line through the origin with a unit slope as given in Figure 8, below, concluding that our fitted Cox-PH model is a perfect fit for the survival times of PTC patients.

**Figure 8:**
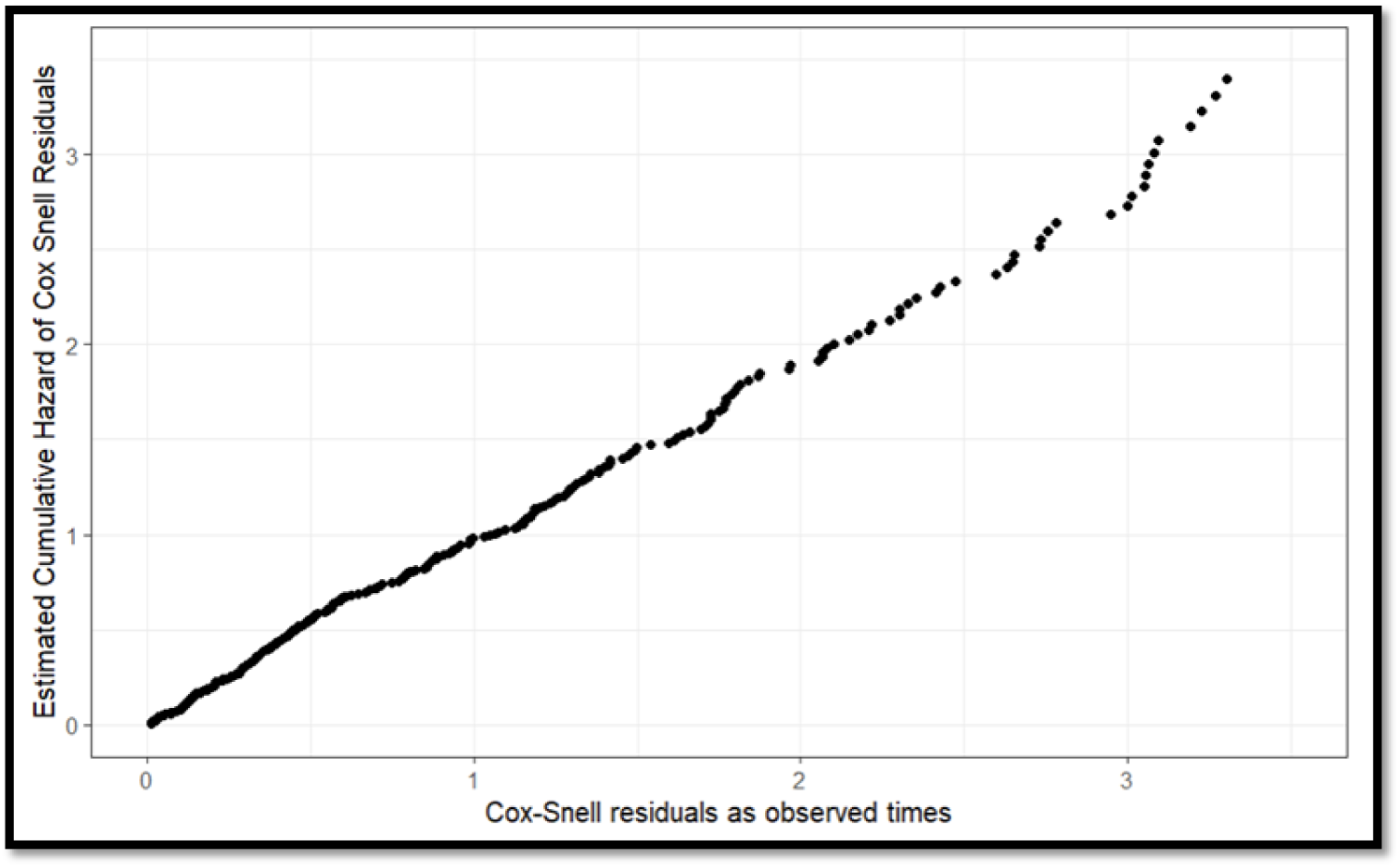
Cox-Snell Residuals Plot for the Proposed Cox-PH Model

## 6 Conclusion

In the present study, we have developed a semi-parametric Cox-PH model for the survival times of PTC patients with five covariates: Age, Malignant tumor size, Sex, Cancer stage, and Race and their two-way interactions to study the association between the hazard ratio and the risk factors. The final Cox-PH model is statistically significant with five individual covariates/ risk factors (but only three covariates are individually statistically significant) and three two-way interaction terms that have been developed in Section 4. In the literature, most of the research studies don’t consider the contribution of the significant interaction terms, which are very important. In our study, we have identified that some individual covariates have HRs greater than 1, but the interaction between those two covariates (for example, between the covariates sex and race) has an HR less than 1. This confirms the importance of including interactions in the Cox-PH model. Moreover, the proposed Cox-PH model satisfies all key assumptions that have been discussed under Section 5 by confirming the accuracy and robustness of our final model. Then, we have ranked the individual covariates and two-way interactions based on the hazard ratio to identify the most influential risk factors.

Thus, the following important results can be summarized for the survival of PTC patients with respect to the risk factors based on our proposed Cox-PH model.

- For the risk factor, Sex, the HR value (1.619503) is greater than 1, which means the risk of death due to PTC is higher with a bad prognosis. That is, a female PTC patient has a higher risk of death than a male patient with PTC.
- For the risk factor, Race, the HR value (2.138914) is greater than 1, which means the risk of death due to PTC is higher with a bad prognosis. That is, a PTC patient from another race has a higher risk of death than a PTC patient from the white.
- For the interaction between Age and Stages, the HR values are greater than 1, which means the risk of death due to PTC is higher with a bad prognosis. That is, a patient with PTC at age between 70 - 96 in the regional or distant stage has a higher risk of death than a patient with PTC at age between 20 - 70 in the localized stage.
- For the interaction between Sex and Race, the HR values are less than 1, which means the risk of death due to PTC is lower with a good prognosis. That is, a female patient of other races has a lower risk of death than a white male patient.
- Based on hazard ratios, the interactions between Age and Stages are the most contributing factors for the death of PTC patients.
- Based on the hazard ratios, the most contributing individual risk factors are Race, Sex, and Malignant tumor size, respectively.

## Data Availability

Surveillance, Epidemiology, and End Results (SEER) program

## 7 Statements and Declarations

### Ethical approval

- We, all authors, Dilmi Abeywardana, Dr. Malinda Iluppangama, and Dr. Chris P. Tsokos have agreed on authorship, read and approved the manuscript, and given consent for submission and subsequent publication of the manuscript. Dilmi Abeywardana acts as the corresponding author for this manuscript.
- We ensure that the manuscript and its contents are not a duplicate publication, and it is completely original.
- We ensure that the manuscript represents our original work and meets the ethical standards set by the Committee on Publication Ethics(COPE).

### Consent to Publish declaration

The authors give full consent to the Pharmaceutical Statistics to publish this manuscript.

### Consent to Participate declaration

Not applicable

### Ethics declaration

Not applicable

### Author Contributions

Conceptualization: Dilmi Abeywardana

Formal analysis: Dilmi Abeywardana and Malinda Iluppangama

Investigation: Dilmi Abeywardana and Malinda Iluppangama

Methodology: Dilmi Abeywardana and Malinda Iluppangama

Writing original draft preparation: Dilmi Abeywardana and Malinda Iluppangama

Writing review and editing: Dilmi Abeywardana Chris Tsokos

Supervision: Chris Tsokos

